# Association of Sleep-Disordered Breathing With Cardiovascular Events and Death in patients With Ischemic Heart Disease: A systematic review and meta-analysis

**DOI:** 10.1101/2021.07.21.21260935

**Authors:** Yaodan Zhang, Jin Tan, Yuyang Miao, Qiang Zhang

## Abstract

**Objective:** Previous studies have identified that sleep-disordered breathing is associated with the increased risk of cardiovascular disease. However, CPAP, the preferred treatment of sleep apnea, does not appear to reduce the risk of adverse cardiovascular events and deaths in ischemic heart disease. Our aim was to determine whether OSA can be a prognostic factor for cardiovascular adverse events and deaths in patients with ischemia heart disease.

**Methods:** We searched Medline via PubMed, Embase, and the Cochrane Library database for clinical studies reporting the major adverse cardiac outcomes of SDB in patients with myocardial ischemia. Sleep apnea tests is mainly performed with cardiorespiratory polygraphy or portable diagnostic device. Summary hazard ratio(HR) and 95% CIs were conducted using random-effects meta-analysis.

**Results:** Eighteen literatures were included, with total participants of 9,138. Sixteen studies reported MACCEs, there was significant association of SDB with MACCES (HR, 1.66[95% CI, 1.41 to 1.94]; P< 0.0001), all-cause mortality (HR, 1.39[95% CI 1.12 to 1.73], P=0.0027) and cardiovascular mortality (HR, 2.12[95% CI 1.23 to 3.65], P=0.007). Subgroup analysis showed that one study was the cause of heterogeneity, while no correlation between SDB and outcomes for sleep apnea severity, follow-up time or whether surgery was performed.

**Conclusion:** SDB is associated with adverse cardiac outcomes in patients with ischemic heart disease, suggesting that sleep apnea can be screened for patients with ischemic heart disease, which can be used as a clinically relevant strategy of secondary prevention to reduce the risk of adverse outcomes.

## 1. Introduction

Ischemic heart disease is an important health burden, and the short - and long-term outcomes have improved following the prompt detection and treatment. But the morbidity, repeat cardiovascular events, and mortality of schemic heart disease remain high. OSA is a common disorder that affects 20-30% of adults and is caused by the collapse of the upper airway during sleep.^1^ And sleep apnea, the recurrent shallow breathing or apnea during sleep, has been shown to have a consistent association with vascular morbidity and mortality.^2, 3, 4^ Studies have shown that the prevalence of OSA varies at different stages after acute coronary syndromes, including pre-operative, post-operative, and recovery or long-term.^5, 6, 7^ A recent meta-analysis found that the incidence of OSA after acute coronary syndromes is high,^8^ but a widespread absence of SDB as a target for screening in patients with ischemic heart disease.

PAP is a preferred treatment for sleep apnea, and has been reported to be associated with a moderate reduction in blood pressure,^9^ sleepiness, quality of life, cardiac ejection fraction, arrhythmias.^10^ Nevertheless, studies based on randomised controlled trial (RCT) has shown that CPAP was not associated with reduced risk of cardiovascular events and death in patients with ACS.^11, 12^ Taken together, these studies seem to suggest that OSA increased cardiovascular risk, but did not appear to increase the poor prognosis in patients with myocardial ischemia. However, a prospective study demonstrated that OSA may be a risk factor for predicting the risk of MACCEs in patients with coronary heart disease after percutaneous coronary intervention (PCI).^13^ And OSA could aggravate ventricular remodeling and decrease the survival rate of mice after myocardial infarction.^14^ Therefore, we aimed to gain insight the correlation between SDB and cardiovascular outcomes in patients with myocardial ischemia, and whether SDB can be used to predict the outcomes of patients with ischemia heart disease, which may provide guidance for early screening and intervention as secondary prevention.

## 2. Methods

### 2.1 Search Strategy

A systematic literature search was done according to the general guidelines outlined in the PRISMA statement.^15^ Medline via PubMed, Embase, and the Cochrane Library database (from their respective inceptions to February 22, 2021) were searched using medical subject headings(MeSH) and relevant text words that consisted of terms relating Sleep Apnea Syndromes and Myocardial Ischemia.(detailed search strategy was shown in appendix 1). There is no language restriction for this literature search.

### 2.2 Trial Inclusion Criteria

observational cohort study of ischemic heart disease patients (including unstable angina, non-ST segment elevation myocardial infarction, ST elevation myocardial infarction) who underwent or not underwent PCI/ coronary artery bypass graft surgery (CABG) that assessed the association of SDB with cardiovascular event were potentially included. In addition, a randomized controlled trial also compared the prevalence of cardiovascular events between ACS patients with OSA who received usual care and ACS patients without OSA.^12^ One study investigated the relationship between moderate-to-severe OSA and major adverse cardiac events in ACS patients who underwent PCI.^16^ Also, one research reported all-cause mortality of severe OSA group in patients with ischemic heart disease or previous myocardial injury.^17^ Duplicate reports, studies with a follow-up duration of less than 12 months, a trial with less than 50 participants were excluded. Researches that did not report on cardiovascular outcomes, populations that have already been diagnosed with SDB at the beginning of the study were also excluded. We also excluded non-English-language studies.

### 2.3 Data Extraction

Two authors independently screened the literature by reading the titles and abstracts according to the inclusion and exclusion criteria to determine the literatures for review in full text. Then, the full-text of each paper was read independently by two authors, and the inclusion of literature was finally determined, and the reports that could not determine whether to be included was determined by a third researcher. After literature screening, the two authors extracted the included article information respectively, including first author, year of publication, nation or region where the study was conducted, inclusion criteria, included study dates, median follow-up duration, number of participants, mean AHI, number/percent of male participants, mean age of participants, mean BMI of participants, number/percent of smoker participants, major adverse cardiovascular or cerebrovascular events (MACCEs), all-cause death, and cardiovascular mortality.

MACCEs, defined as a composite of cardiac death, nonfatal myocardial infarction, nonfatal stroke, and unplanned revascularization, or hospital admission for congestive heart failure or unstable angina. All-cause mortality, cardiovascular mortality, myocardial infarction, stroke, ischemic heart disease hospitalization, all repeat revascularization were also extracted. These outcome events were evaluated based on reported HR.

### 2.4 Quality assessment

The Newcastle-Ottawa Scale was used to assess the quality of observational cohort studies. Based on the previous work, we defined ≥7 scores as high quality, 4-6 scores as medium quality, and <4 scores as low quality. The authors independently evaluated the quality of each study and resolved the discrepancies through discussion.

### 2.5 analysis

Data analysis was performed using R (version 4.0.3), and the Pooled outcome measures were combined using a random effects model. Heterogeneity was estimated by Cochran Q statistic, and P<0.05 was considered to be heterogeneous. I^2^ values used to estimate the percentage of heterogeneity with values 0–25%, 25%-75%, more than 75% correspond to low, moderate, and high probability of differences. Subgroup analysis were done to determine the source of heterogeneity according to whether surgery had been performed in the included studies, OSA severity and the duration of follow-up. In addition, Egger regression was conducted to evaluate the funnel plot for asymmetry, indicating the evidence of publication bias.

## 3. Results

### 3.1 Search Results and Characteristics of Included Studies

The literature search identified 7908 publications, among which, 179 literatures were reviewed in full text after de-duplication and reading the title/abstract. Finally, 18 literatures were included by reading the full text, including 12 prospective cohort studies, 3 retrospective studies, 2 ambiguous studies, and 1 randomized controlled trial. Fig 1 is the detailed information of the literature screening. Table1 summarized the detailed characteristics of included studies. The number of participants of each study ranged from 68 to 1311. Median follow-up time for all studies was 30.5 months, and the diagnosis of sleep apnea was based on conventional polysomnographic or portable diagnostic device in all but 1 trial that used a Nocturnal pulse oximetry^18^. The detailed outcomes of participants of the included literatures were shown in Table 2. Newcastle–Ottawa Scale was used to evaluate the quality of research, which was shown in appendix 2.

**Table 1.** Patient demographics of included studies.

| Study | Number of patients | | AHI (events per hour) | | Males sex, n (%) | | Mean ± SD or median (range) | | Mean BMI ± SD (kg/m<sup>2</sup>) | | Smoker, n (%) | |
| --- | --- | --- | --- | --- | --- | --- | --- | --- | --- | --- | --- | --- |
| | | | Mean ±SD or median (IQR) | | | | age (years) | | | | | |
|  | SDB | NO-SDB | SDB | NO-SDB | SDB | NO-SDB | SDB | NO-SDB | SDB | NO-SDB | SDB | NO-SDB |
| Fan et.al | 403 | 401 | 31.2(21.8–43.8) | 7.4 (3.8–10.6) | 342(84.9) | 322(80.3) | 57.7±10.2 | 57.2±10.2 | 27.7±3.5 | 25.8±3.4 | - | - |
| He et al. | 158 | 425 | 29.0±12.2 | 5.3±4.2 | 96(60.8) | 157(36.9) | 64.5 ± 12.9 | 63.4±12.7 | 24.9±3.6 | 23.7±3.8 | 52(32.9) | 97(22.8) |
| Koo et al. | 513 | 494 | 30.1(20.6–44.7) | 5.85(3.0–10.23) | 442(86.2) | 429(86.8) | 62(56–68) | 61(56–67) | - | - | 139(27.1) | 155(31.4) |
| Lee et al. | 594 | 717 | 28.9(21.1) | 5.9(7.1) | 523(88.1) | 594(82.9) | 59.0(10.2) | 57.5(10.3) | 26.6(3.9) | 25.0 (3.4) | 210(35.4) | 255(35.6) |
| Lin et al | 93 | 465 | - | - | 79(85) | 385(83) | 63±12 | 64±13 | - | - | - | - |
| liu et al. | 89 | 109 | - | - | 61 (68.5) | 71(65.1) | 61.45±7.30 | 61.57 ± 7.25 | 28.8±4.5 | 24.6±3.1 | - | - |
| Loo et al. | 24 | 44 | 24.0(16.9–52.0) | 4.9 (0.3–14.9) | 18(75.0) | 41(93.2) | 56.7(5.6) | 52.8(10.0) | 27.5(3.9) | 24.4(3.3) | 4(16.7) | 28 (63.6) |
| Mazaki et al. | 126 | 115 | 13.5(15.3) | 2.1(2.2) | 69(78) | 66 (75) | 63±13 | 63±12 | 24.7±3.8 | 24.4±3.5 | - | - |
| Moore et al. | 133 | 259 | - | - | - | - | - | - | - | - | - | - |
| Nakashima et al. | 124 | 148 | - | - | 96 (77) | 108 (73) | 71(61–78) | 65(56–75) | - | - | - | - |
| Sánchez-de-la-Torre et al. | 626 | 596 | 35.5(18.3) | - | 530 (85) | - | 60.7(10%) | - | 29.4(4.29) | - | 291(47) | - |
| Valham et al. | 211 | 181 | - | - | 73(35) | 60(33) | 60.7±7.0 | 59.3±7.8 | 27.3±3.7 | 26.8±3.3 | - | - |
| Won et al. | 130 | 151 | 51±17 | 15±11 | 127(97.7) | 147(97.3) | 65.6±11 | 64.6±11 | 35.4±8 | 32.9±8 | - | - |
| Yatsu et al. | 296 | 243 | - | - | 259 (87.5) | 196(80.7) | 68.2±11.5 | 66.2 ± 10.7 | 25.5±3.9 | 23.7±3.1 | - | - |
| Zeng et al. | 373 | 379 | 33.7±15.1 | 7.1±4.0 | 315(84.5) | 306(80.7) | 57.6 ± 10.4 | 56.8 ± 10.0 | 27.6 ± 3.4 | 25.8 ± 3.3 | - | - |
| Zhang et al. | 152 | 188 | 30.9 (21.8, 43.5) | 8.4 (3.9, 11.9) | 120 (78.9) | 129 (68.6) | 65.6±10.3 | 64.0±10.4 | 25.4±2.9 | 24.5±3.1 | - | - |
| Leão et al. | 46 | 27 | 30.6 ± 23.0 | 2.3 ± 3.2 | 38 (83%) | 17 (63%) | 63.5 ± 10.3 | 60.6 ± 12.7 | 27.8 ± 3.8 | 27.2 ± 3.4 | 8 (17%) | 5 (18%) |
| Lee et al. | 44 | 61 | - | - | 43 (97.7) | 60 (98.4) | 55.2 (11.0) | 50.9 (8.5) | 26.1 (3.4) | 24.0 (2.9) | 24 (54.6) | 37 (60.7) |

IQR, interquartile range

**Table 2.**
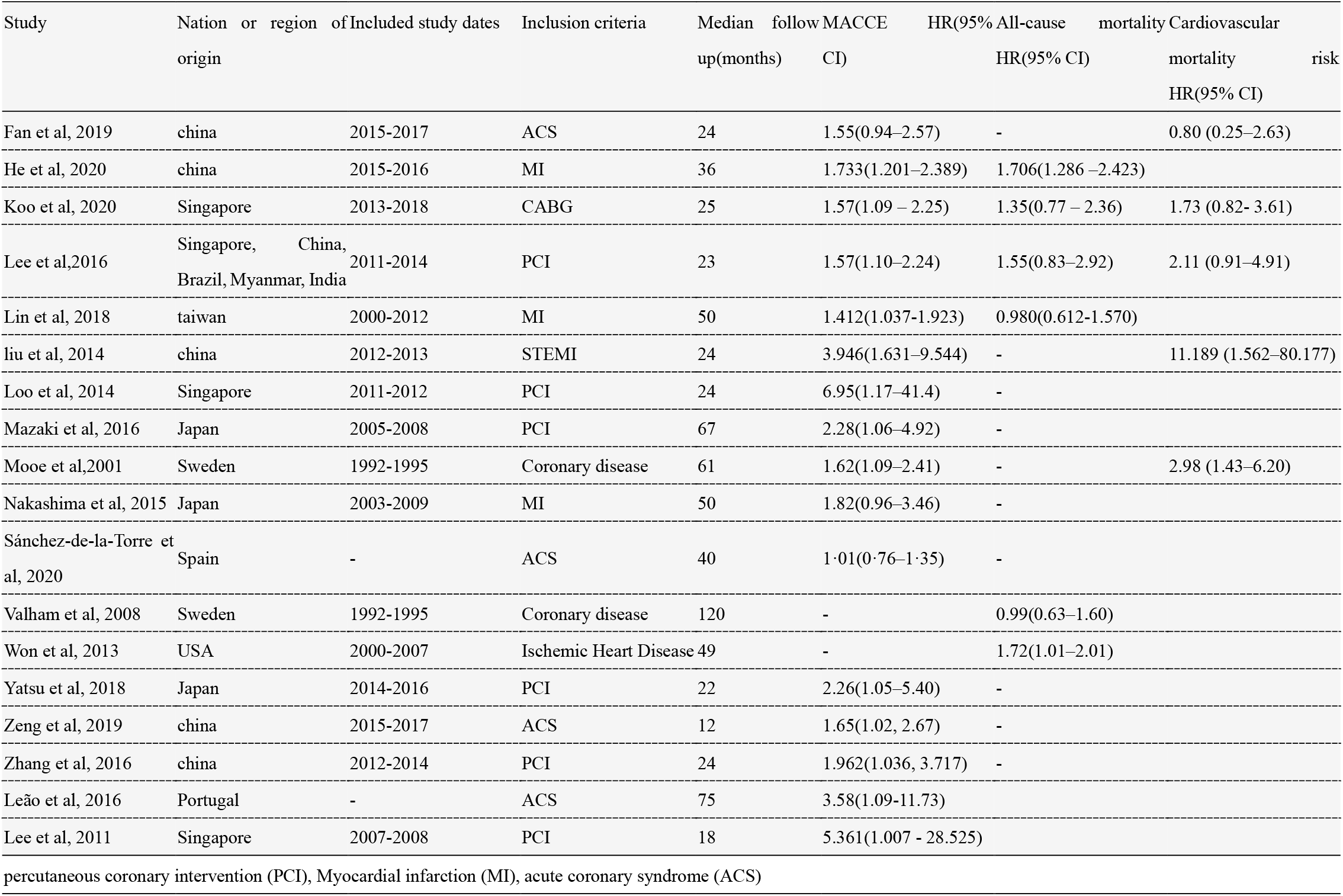
Outcomes of included studies assessing the effect of sleep-disordered breathing on the prognosis of ischemia heart disease.

**Fig 1.**
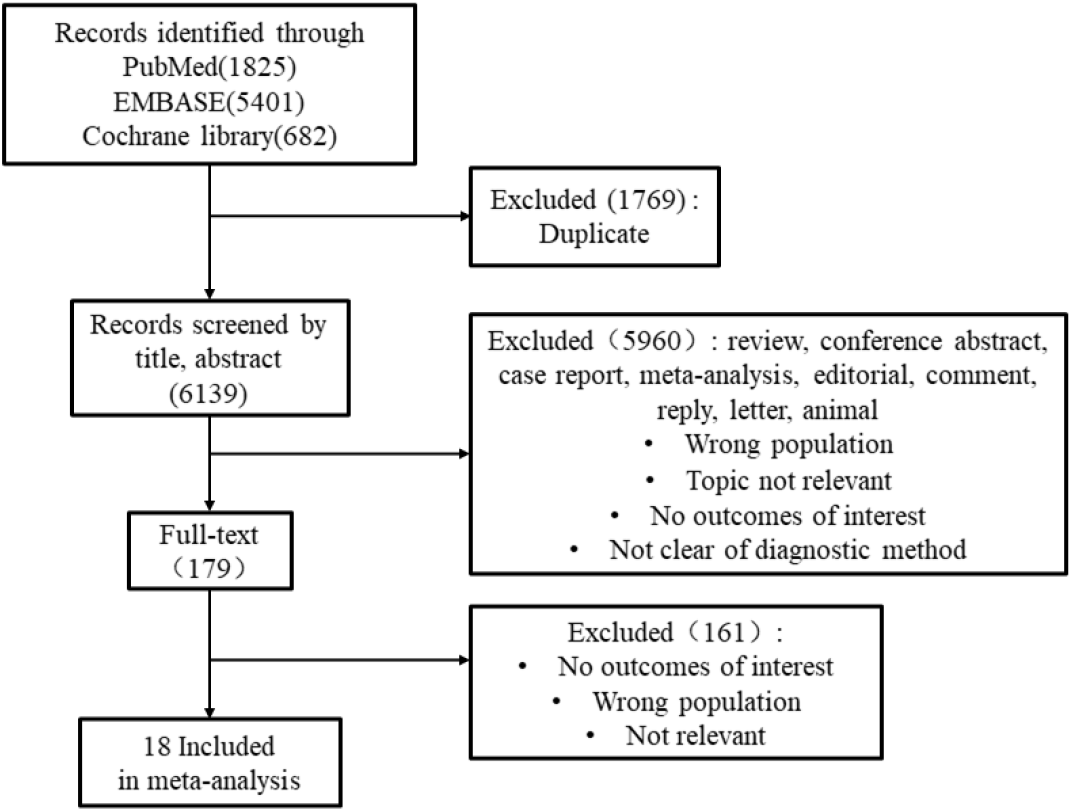
Detailed flow chart of literature search.

### 3.2 Major adverse cardiovascular or cerebrovascular events (MACCEs)

Sixteen studies reported an increased Hazard ratio (HR) of MACCEs outcomes with SDB compared with controls in patients with myocardial ischemia,^12,13,16,18,19,20,21,22,23,24,25,26,27,28,29,30^ with the combined HR of 1.66(95% CI 1.41-1.94, P< 0.0001) (Fig 2). Some evidence of statistical heterogeneity (Cochran Q = 23.00, p=0.08; I^2^ = 35%) was found by pooled effect size, which might be in part attributable to 1 randomized controlled trial. Subgroup analysis found no significant associations of SDB with reported relative risk of MACCEs for different levels of SDB severity, follow-up time, whether operate on myocardial ischemia. (Table 3) There was some evidence of bias (Egger intercept = -0.0631, p < 0.0001) (Fig 5).

**Table 3.** Association of sleep-disordered breathing with cardiovascular outcomes and deaths in trial subgroups.

| Subgroup | Number of studies | HR(95%CI) | Heterogeneity |
| --- | --- | --- | --- |
| All studies | 16 | 1.66(1.41-1.94) | I<sup>2</sup> =35%, $\tau^2$=0.0323, p=0.08 |
| Degree | 9 | 1.56(1.27-1.92) | I<sup>2</sup> =42%, $\tau^2$=0.0379, p=0.09 |
| severe | 6 | 1.50(1.14-1.96) | I<sup>2</sup> =47%, $\tau^2$=0.0491, p=0.09 |
| moderate | 3 | 1.72(1.27-2.33) | I<sup>2</sup> =23%, $\tau^2$=0.0181, p=0.27 |
| **Median_follow_up** |  |  |  |
| 1-2year | 8 | 1.89(1.48-2.41) | I<sup>2</sup> =15%, $\tau^2$=0.0190, p=0.31 |
| more than 2years | 8 | 1.52(1.25-1.85) | I<sup>2</sup> =40%, $\tau^2$=0.0296, p=0.11 |
| Study_types | 14 | 1.64(1.38-1.95) | I<sup>2</sup> =40%, $\tau^2$=0.0381, p=0.06 |
| cohort study | 13 | 1.68(1.47-1.92) | I<sup>2</sup> =0%, $\tau^2$=0, p=0.46 |
| randomised controlled trial | 1 | 1.01(0.76-1.35) | - |
| **Surgery** |  |  |  |
| no | 9 | 1.58(1.28-1.96) | I<sup>2</sup> =48%, $\tau^2$=0.0458, p=0.05 |
| yes | 7 | 1.77(1.43-2.19) | I<sup>2</sup> =0%, $\tau^2$=0, p=0.46 |

**Fig 2.**
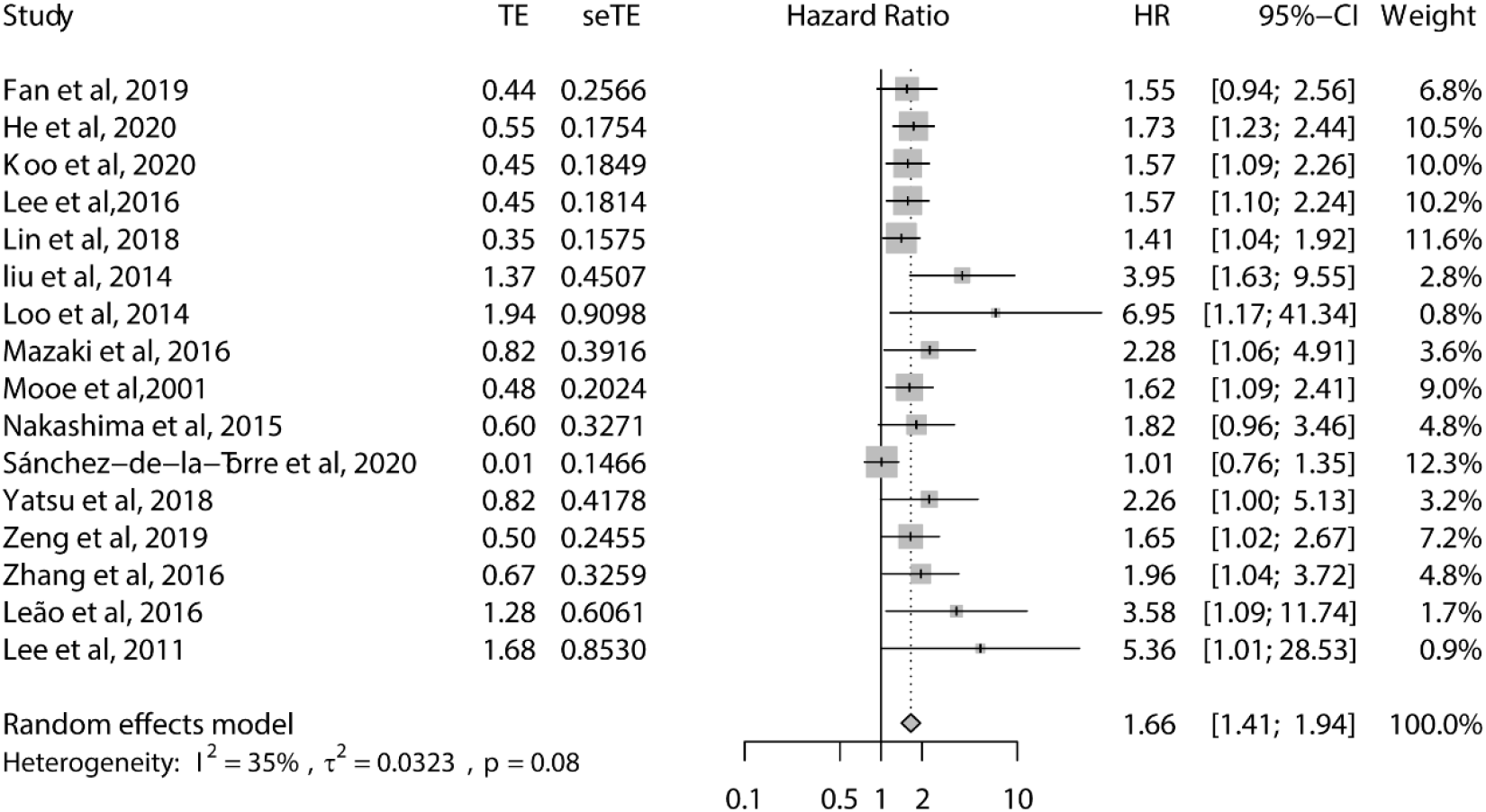
Forest plot of the association of sleep-disordered breathing with major adverse cardiovascular or cerebrovascular events and deaths.(pooled hazard ratio 1.66, 95% CI 1.41 to 1.94, P< 0.0001)

### 3.3 All-cause mortality

Six studies reported an increased Hazard ratio (HR) of all-cause mortality with SDB compared with controls in patients with myocardial ischemia,^13, 17, 26, 29, 30, 31^ with the pooled HR of 1.39(95% CI 1.12-1.73, p=0.0027) (Fig 3). There was no obvious statistical heterogeneity (Cochran Q = 7.27, p =0.2; I^2^ = 31%), and no evidence of bias (Egger intercept = 0.8883, p = 0.2548).

**Fig 3.**
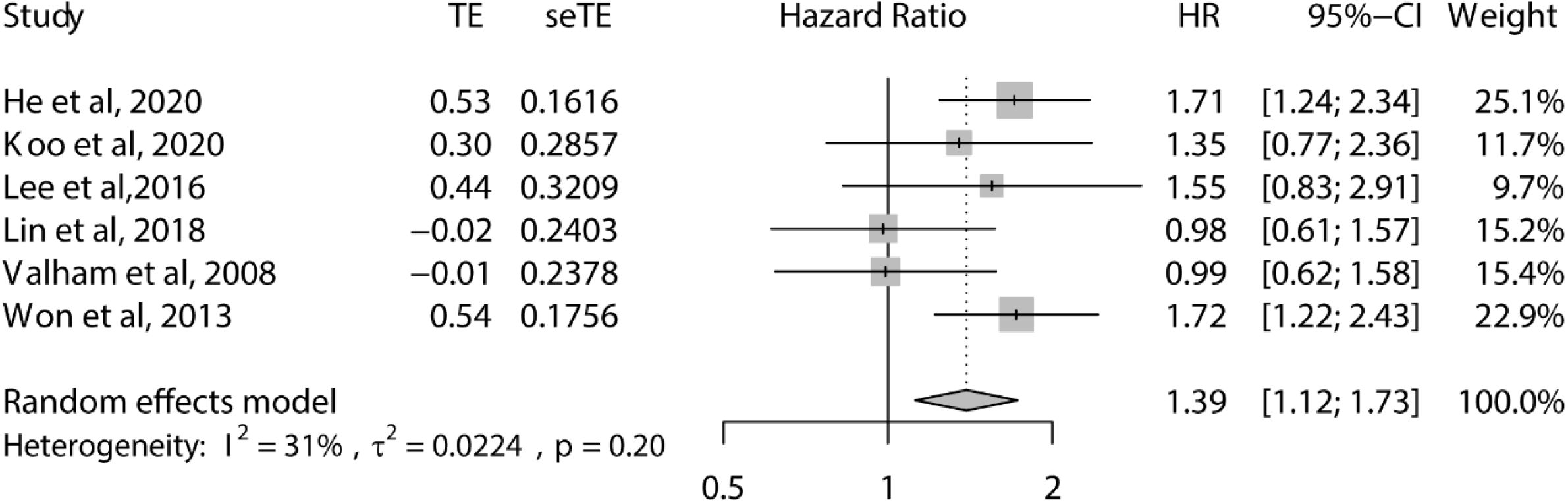
Forest plot of the association of sleep-disordered breathing with all-cause mortality (pooled hazard ratio 1.39, 95% CI 1.12 to 1.73, P=0.0027).

### 3.4 Cardiovascular mortality

Five studies reported the increased Hazard ratio (HR) of cardiovascular mortality,^13, 19, 21, 27, 30^ with the pooled HR of 2.12(95% CI 1.23-3.65, p=0.007) (Fig4). There was no significant evidence of statistical heterogeneity (Cochran Q = 6.49, p =0.165; I^2^ = 38.4%), and no evidence of bias (Egger intercept = 0.3260, p = 0.6848).

**Fig 4.**
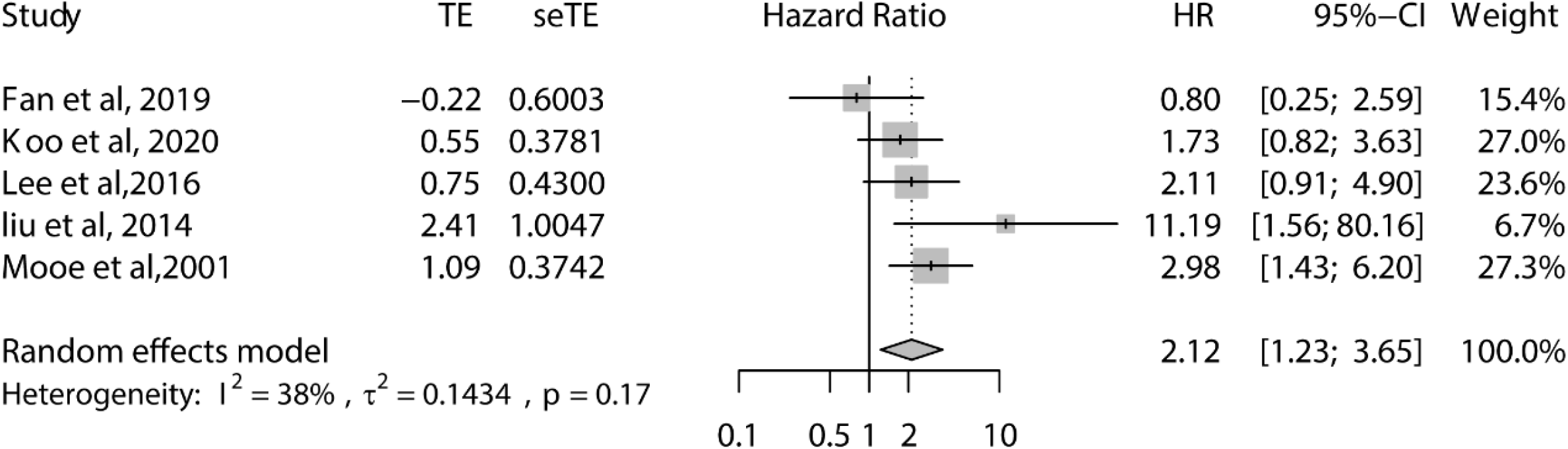
Forest plot of the association of sleep-disordered breathing with cardiovascular mortality (pooled hazard ratio 2.12, 95% CI 1.23 to 3.65, P=0.007).

**Fig 5.**
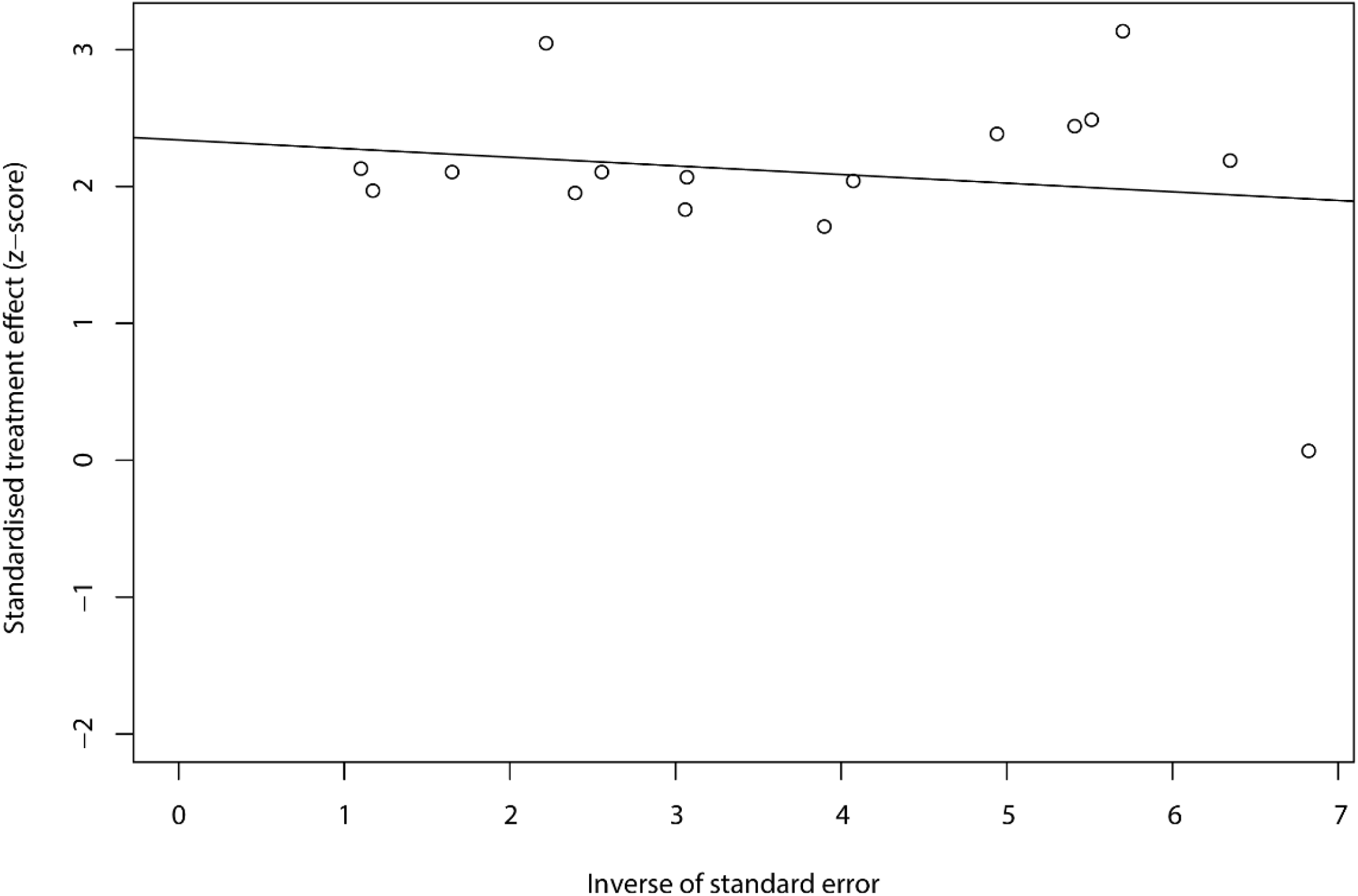
The Egger Funnel plot of included studies’ effect sizes on major adverse cardiovascular or cerebrovascular events and deaths.

## 4. Discussion

These results suggest that SDB is relevant to the increased major adverse cardiovascular or cerebrovascular events in patients with myocardial ischemia. This is consistent with an observational study showing that sleep apnea is associated with the increase in the risk of cardiovascular events.^32^ In addition, in 2017, a meta-analysis showed that OSA was closely associated with a rising risk of cardiovascular events in patients underwent PCI.^33^ OSA also was an important risk factor for major adverse cardiac or cerebrovascular events in patients undergoing cardiac surgery.^34^ Our results show that OSA increases adverse cardiovascular outcomes in patients with myocardial ischemia at large and not just limited to patients undergoing surgery (PCI/CABG).

The present study also shows that SDB is associated with all-cause mortality and cardiovascular death risk in patients with myocardial ischemia, and the increase of cardiovascular mortality may be related to the comorbidities of cardiovascular disease including hypertension, diabetes and dyslipidaemia. A meta-analysis concluded that bariatric surgery can reduce all cause long-term mortality, cardiovascular mortality and development of ischemic heart disease to some extent.^35^ We all known that obesity is an important risk factor for OSA, and our study cannot rule out the influence of obesity-related diseases on adverse cardiovascular outcomes.

A small sample of study suggested that CPAP treatment could alleviate MACCEs.^26^ Nevertheless, a large meta-analysis of CPAP, the preferred treatment for sleep apnea, has shown that it does not reduce the risk of cardiovascular outcomes.^11^ At the same time, a meta-analysis indicated that adequate CPAP treatment (defined as at least 4 h/d) compared with usual care alone have an positive impact on the MACCEs in patients with OSA, but stratified analysis showed that CPAP use is associated with the reduction of cerebrovascular adverse events and stroke, not cardiac outcomes.^36^ The different effects of CPAP treatment on cardiac outcomes and cerebrovascular systems are consistent with a new result of an increased incidence of cerebrovascular circulation and stroke in older patients with sleep apnea, but not the prevalence of coronary heart disease.^37^ In 2020, a randomized controlled study showed that OSA with usual care alone compared with patients without OSA was not associated with the prevalence of cardiovascular events in patients with ACS, and CPAP treatment did not reduce the prevalence of cardiovascular outcomes or death.^12^ Although CPAP therapy may have other benefits, the present findings suggest that CPAP treatment is not supported for the prevention of the adverse cardiac outcomes. Thus, more large, prospective, multicenter studies are needed to further validate the association between SDB and MACCEs in patients with ischemic heart disease in the future, especially to distinguish cardiac and cerebrovascular adverse events.

However, our study showed that SDB increased MACCEs, all-cause mortality and cardiovascular mortality, but not distinguish between cardiac and cerebrovascular adverse events. After excluding this randomized controlled trial, the forest plot results showed no statistical heterogeneity by the pooled effect size. Studies have shown a decrease in in-hospital mortality in patients with ST elevation myocardial infarction with recognized OSA compared with patients without OSA, but there was no difference in incidence of in-hospital cardiac arrest.^38^ Shah et al. showed that OSA patients with acute MI have fewer severe cardiac compared to patients without OSA, possibly due to the cardioprotective effect of sleep apnea during acute myocardial infarction via ischemic preconditioning.^39^ Also, Chang et al. have shown that intermittent hypoxic (IH) preconditioning can improve the heart injury caused by ischemia/reperfusion.^40^ However, Du et al. demonstrated that MI+CIH OSA can aggravate cardiac remodeling and decrease the survival rate of mice.^14^ Hence, whether SDB occurs before or after myocardial ischemia may have a certain influence on the outcomes.

Wu et al. found that untreated moderate to severe OSA had an increased risk of repeat revascularization compared with the OSA treated with CPAP group, but no significant in MACCEs and MACEs.^41^ Two studies compared the major adverse events of severe OSA to non-severe OSA in patients with myocardial ischemia, which showed a negative prognostic impact.^17, 20^ Therefore, there may be a dose-response relationship between the severity of OSA and the adverse prognostic outcomes of CHD.

The limitations of our study: the studies we included did not indicate whether OSA patients were treated with CPAP, and patients with other complications or non-first visit for CHD, which could have some impact on outcomes, but the included literature did not clearly indicate this. And the diagnostic criteria for sleep apnea of part literatures are different, with sleep apnea was defined as an AHI ≥15 events/h, or was defined as an AHI ≥5 events/h, or even was defined as an AHI ≥10 events/h, as well as the severity of ischemic heart disease and whether undergoing surgery (PCI/CABG), not distinguish between OSA and CSA, which may be the possible causes of heterogeneity. In addition, the sleep studies were conducted with polysomnography or portable monitoring device, and questionnaires and a home-based sleep study were excluded, so our study may not be widely representative of the population.

One limitation of our study was that the HR of the included literatures was mostly adjusted, while some literatures were not adjusted, but it was difficult to be further unified. In addition, the screen of SDB into our included studies of patients with ischemic heart disease has proved difficult in some cases, particularly when the sleep apnea was diagnosed in an untimely manner. Patients diagnosed with SDB after IHD were selected, but it is difficult to determine whether these patients were previously free of SDB, especially when the latency between coronary heart disease and sleep apnea tests varied significantly in one study, which may account for some of the differences in clinical outcomes. Moreover, there was statistical evidence for publication bias, and part of the single-center study may have a certain degree of bias. Some negative results were not published, which could be the partly possible reason for our results. Therefore, future studies are needed to verify the conclusion.

## 5. Conclusion

Our results indicate that SDB was associated with the risk of MACCEs, all-cause mortality, and cardiovascular mortality in patients with ischemic heart disease, but the distinguish of cardiac and cerebrovascular adverse events needs to be further studied. Routine screening of sleep apnea in patients with myocardial ischemia may serve as a secondary prevention strategy to reduce the risk of major adverse cardiovascular and cerebrovascular outcomes. The further studies may best be achieved through increased collaboration among the cardiology, sleep medicine, and clinical trial communities. Also, more high-quality and rigorous researches are needed to avoid publication bias and validate the results.

## Data Availability

Not Applicable.

## Acknowledgements

This research was funded by National Natural Science Foundation of China (Grant No.81970085 and 81670086), the Tianjin Science and Technology Plan Project (Grant No. 17ZXMFSY00080).

## Author contributions

Concept and design: Yaodan Zhang, Qiang Zhang, Jin Tan. Acquisition, analysis, or interpretation of data: Yaodan Zhang, Jin Tan, Qiang Zhang. Drafting of the manuscript: Yaodan Zhang. Software and Statistical analysis: Yaodan Zhang. Language modification and guidance: Yuyang Miao. All authors confirmed the final version of the manuscript for submission.

## Ethics statement

Not Applicable.

## Search strategy

**Appendix 1.**
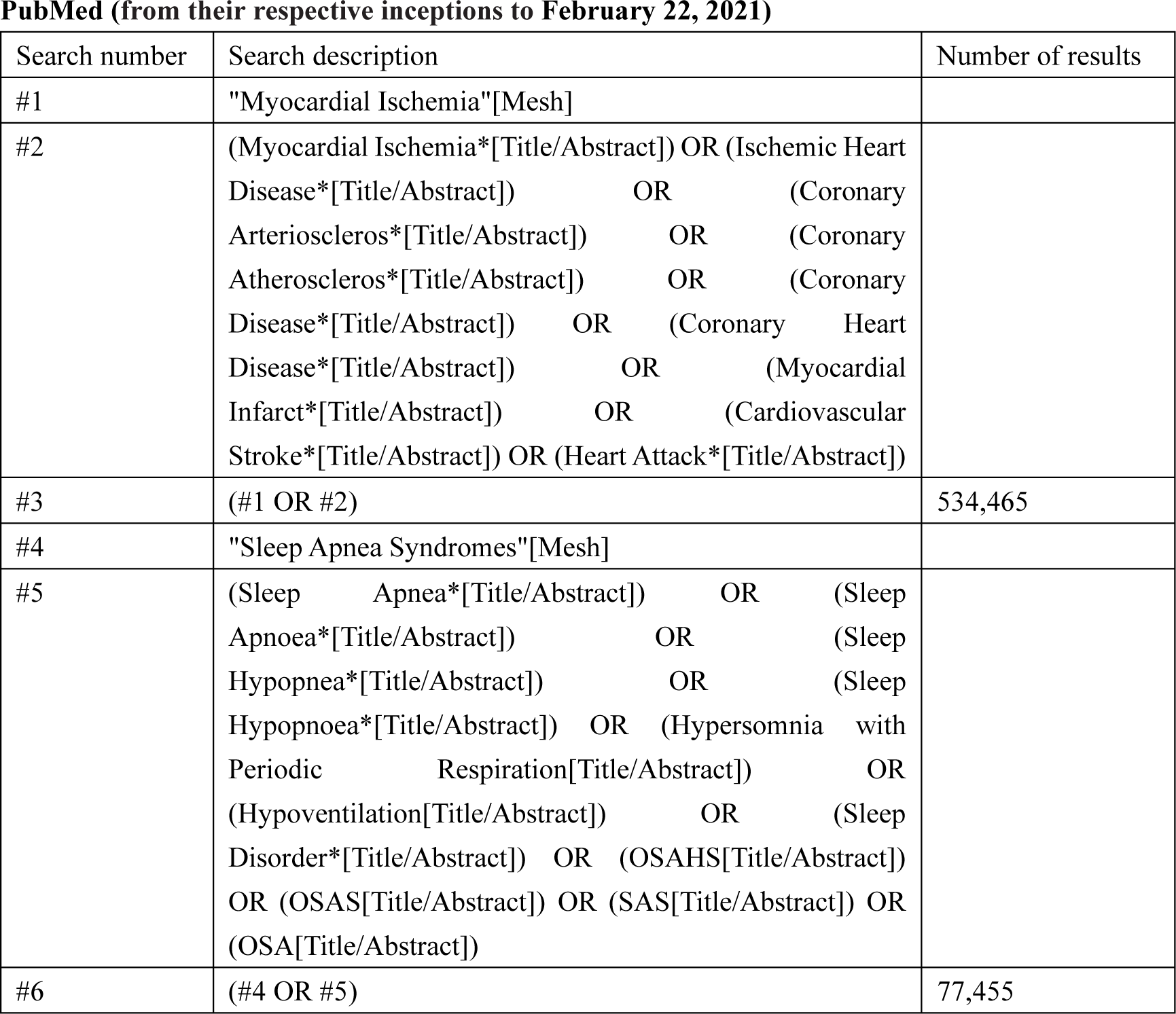
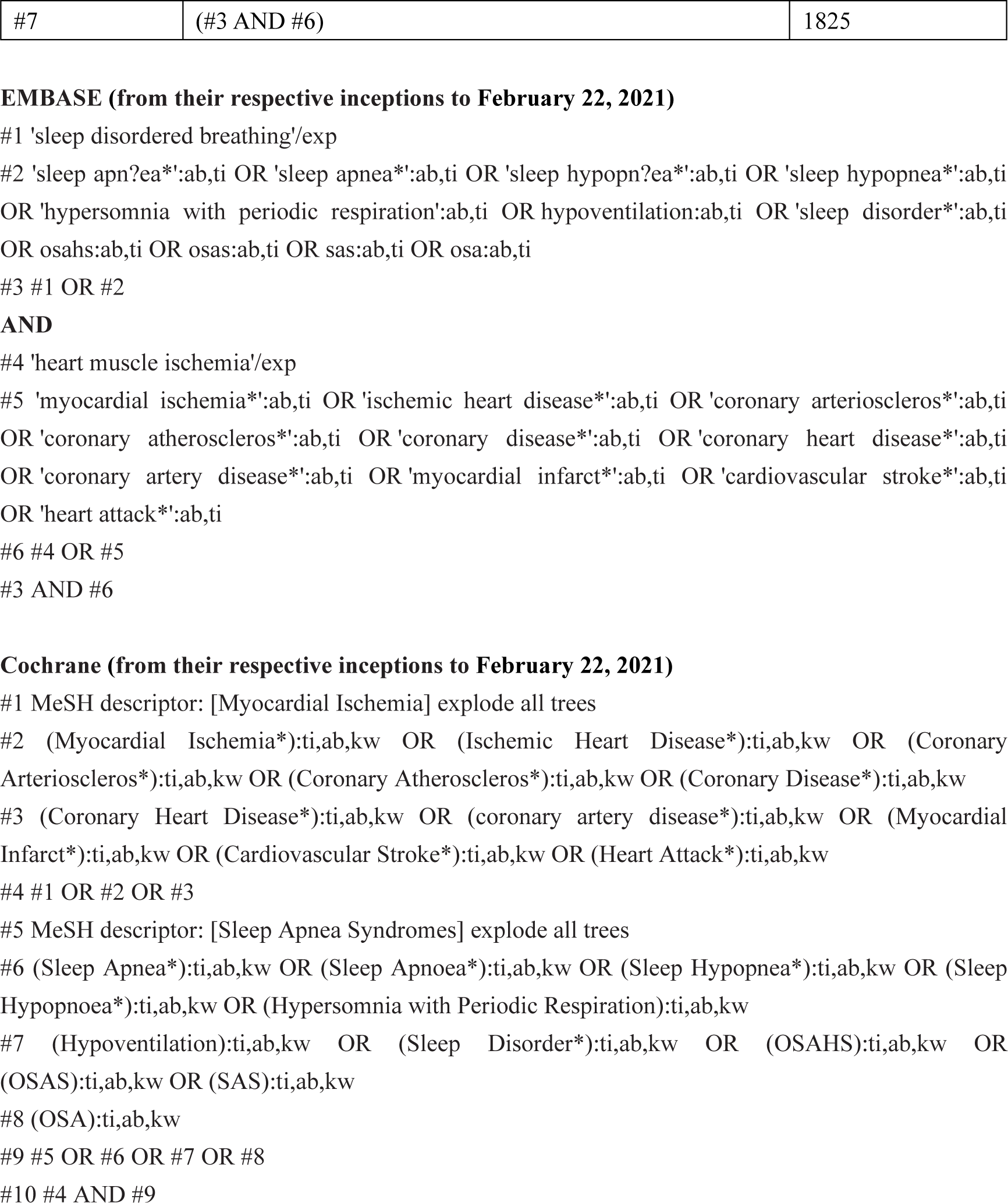
Search strategy

**Appendix 2.**
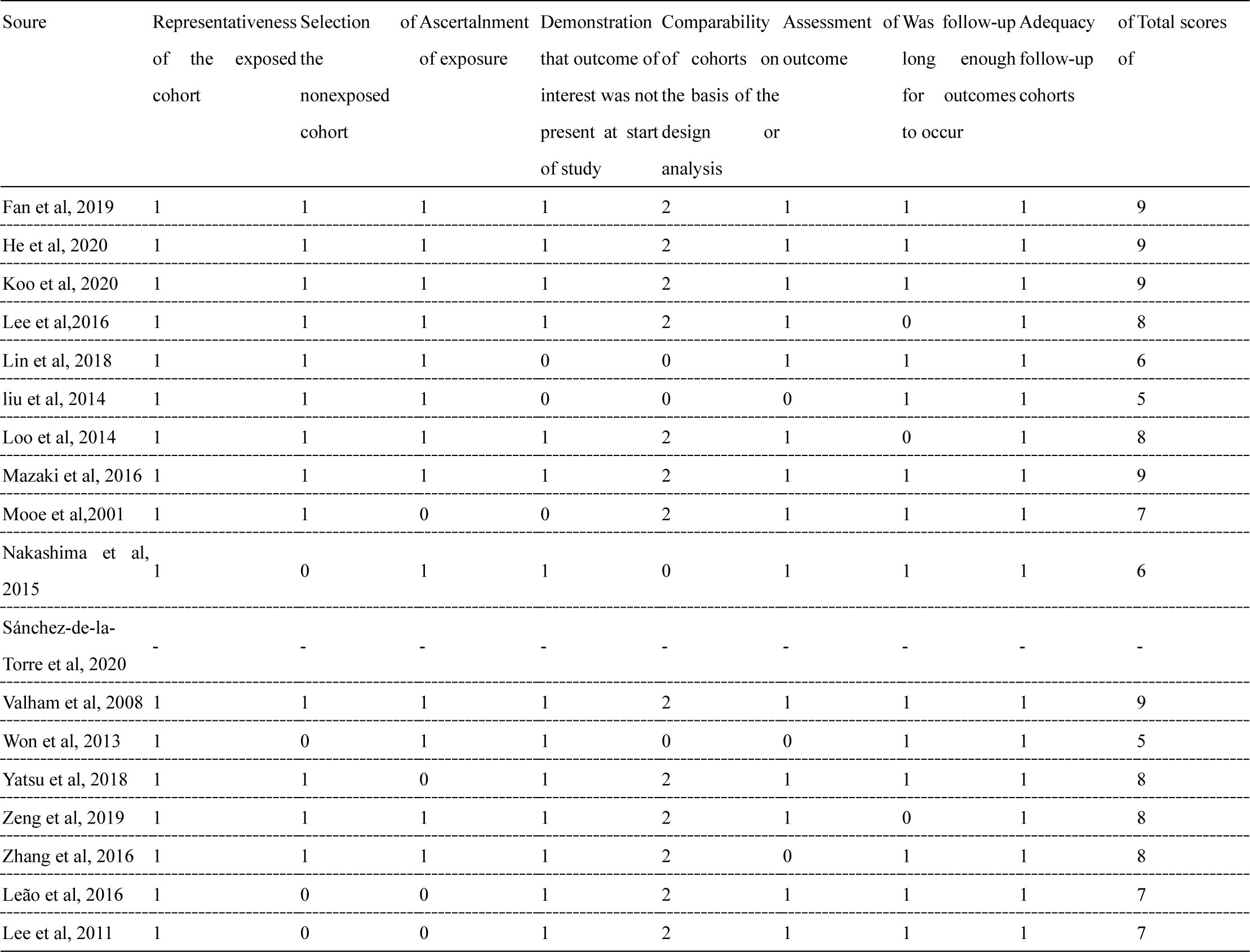
Quality assessment (Newcastle-Ottawa Scale).

