## Supplementary material for "Association of Sleep-Disordered Breathing With Cardiovascular Events and Death in patients With Ischemic Heart Disease: A systematic review and meta-analysis": cover letter (from another version of this preprint: version 1, 2021-07-23)

Dear Editors,

We would like to submit the enclosed manuscript entitled “Association of Sleep-Disordered Breathing With Cardiovascular Events and Death in patients With Ischemic Heart Disease： A systematic review and meta-analysis”, which we wish to be considered for publication in “BMJ global health”. We submit this manuscript based on the following reasons:

Sleep apnea is an important cardiovascular risk factor, with an increased prevalence in patients with myocardial ischemia. Some studies have shown that sleep apnea is a risk factor affecting the prognosis of patients with myocardial ischemia. However, a recent meta-analysis showed that CPAP, the preferred treatment for sleep apnea, did not reduce adverse cardiovascular events and deaths. One study showed that OSA increased adverse cardiovascular outcomes in patients with PCI. This led us to consider whether sleep apnea is positively associated with poor cardiovascular outcomes in patients with myocardial ischemia? We proved that SDB can indeed increase the risk of cardiovascular and cerebrovascular adverse events and deaths, all-cause mortality and cardiovascular mortality in patients with ischemia heart disease, which may provide reference for subsequent routine screening of sleep apnea and early intervention in patients with ischemia heart disease. We believe this manuscript is valuable for all the clinician who are focus on the sleep-disordered breathing and cardiovascular outcomes, and can provide guidance for hospital-community management of patients with coronary heart disease.

No conflict of interest exits in the submission of this manuscript, and manuscript is approved by all authors for publication. I would like to declare on behalf of my co-authors that the work described was original research that has not been published previously, and is not currently being considered for publication elsewhere, in whole or in part. All the authors listed have approved the manuscript that is enclosed.

We deeply appreciate your consideration of our manuscript, and we look forward to receiving comments from the reviewers. If you have any queries, please don’t hesitate to contact me at the address below.

Thank you and best regards.

Yours sincerely,

Qiang Zhang

Yaodan Zhang

**Details of corresponding author:**

Full name: Qiang Zhang.

Affiliation: Department of Geriatrics, Tianjin Medical University General Hospital; Tianjin Geriatrics Institute.

Full postal address: Anshan Road NO.154, Tianjin 300052, China

| Study | TE | seTE | HR | 95%−CI | Weight |
|---|---|---|---|---|---|
| Fan et al, 2019 | 0.44 | 0.2566 | 1.55 | [0.94; 2.56] | 6.8% |
| He et al, 2020 | 0.55 | 0.1754 | 1.73 | [1.23; 2.44] | 10.5% |
| Koo et al, 2020 | 0.45 | 0.1849 | 1.57 | [1.09; 2.26] | 10.0% |
| Lee et al,2016 | 0.45 | 0.1814 | 1.57 | [1.10; 2.24] | 10.2% |
| Lin et al, 2018 | 0.35 | 0.1575 | 1.41 | [1.04; 1.92] | 11.6% |
| liu et al, 2014 | 1.37 | 0.4507 | 3.95 | [1.63; 9.55] | 2.8% |
| Loo et al, 2014 | 1.94 | 0.9098 | 6.95 | [1.17; 41.34] | 0.8% |
| Mazaki et al, 2016 | 0.82 | 0.3916 | 2.28 | [1.06; 4.91] | 3.6% |
| Mooe et al,2001 | 0.48 | 0.2024 | 1.62 | [1.09; 2.41] | 9.0% |
| Nakashima et al, 2015 | 0.60 | 0.3271 | 1.82 | [0.96; 3.46] | 4.8% |
| Sánchez−de−la−Torre et al, 2020 | 0.01 | 0.1466 | 1.01 | [0.76; 1.35] | 12.3% |
| Yatsu et al, 2018 | 0.82 | 0.4178 | 2.26 | [1.00; 5.13] | 3.2% |
| Zeng et al, 2019 | 0.50 | 0.2455 | 1.65 | [1.02; 2.67] | 7.2% |
| Zhang et al, 2016 | 0.67 | 0.3259 | 1.96 | [1.04; 3.72] | 4.8% |
| Leão et al, 2016 | 1.28 | 0.6061 | 3.58 | [1.09; 11.74] | 1.7% |
| Lee et al, 2011 | 1.68 | 0.8530 | 5.36 | [1.01; 28.53] | 0.9% |
| Random effects model | | | 1.66 | [1.41; 1.94] | 100.0% |

Heterogeneity: $I^2 = 35\%$, $\tau^2 = 0.0323$, $p = 0.08$

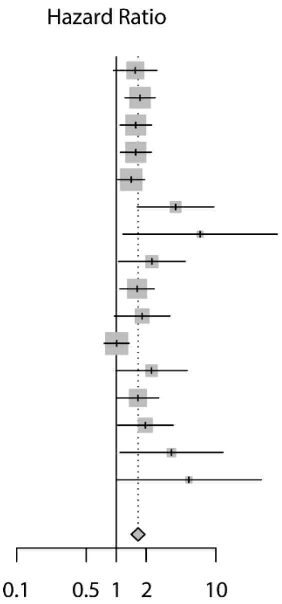
